# Performance of popular pulse oximeters compared with simultaneous arterial oxygen saturation or clinical-grade pulse oximetry: a cross-sectional validation study in intensive care patients

**DOI:** 10.1101/2021.03.09.21253181

**Authors:** Ralf E. Harskamp, Luuk Bekker, Jelle C.L. Himmelreich, Lukas De Clercq, Evert P.M. Karregat, Mengalvio E. Sleeswijk, Wim A.M. Lucassen

**Affiliations:** Amsterdam UMC, University of Amsterdam, Department of General Practice, Amsterdam Cardiovascular Sciences and Amsterdam Public Health Research Institutes, Amsterdam, the Netherlands; Flevoziekenhuis, Department of Intensive Care medicine, Almere, the Netherlands

**Keywords:** community, oxygen, hypoxemia, pulse oximeter

## Abstract

**Background:** The demand for pulse oximeters is high in the current COVID-19 pandemic. Despite their popularity, clinical studies to evaluate the reliability in obtaining information on a patient’s oxygenation status, are lacking.

**Aim:** To evaluate the performance of pulse oximeters under clinical conditions, with arterial blood gas measurement (SaO_2_) as reference standard.

**Methods:** We studied the accuracy of ten top-selling pulse oximeters in Europe and the USA in an intensive care population at the Flevoziekenhuis in Almere, the Netherlands. The studied pulse oximeters were: AFAC FS10D, AGPTEK FS10C, ANAPULSE ANP 100, Cocobear, Contec CMS50D1, HYLOGY MD-H37, Mommed YM101, PRCMISEMED F4PRO, PULOX PO-200, and Zacurate Pro Series 500DL. Adult patients in whom an SaO2 measurement was obtained as part of routine care were asked for inclusion. Directly after obtaining the SaO2 blood sample we obtained the pulse oximeter readings (SpO_2_) of the investigational devices, in random order. Outcomes were bias (SpO_2_ – SaO_2_) mean, root mean square difference (A_RMS_), mean absolute error (MAE), and accuracy in identifying hypoxemia (SaO_2_≤90%). As a clinical index test, we included a hospital-grade SpO_2_-monitor.

**Results:** In 35 consecutive patients we obtained 2,258 SpO_2_-readings and 234 SaO_2_-samples. Mean bias ranged from −0.6 to −4.8. None of the pulse oximeters had an A_RMS_≤3%, the requirement set by ISO-standards and required for FDA 501(k) clearance. The MAE ranged from 2.3 to 5.1, and 5 out of 10 pulse oximeters met the requirements of ≤3%. For hypoxemia, negative predictive values were 98-99%. Positive predictive values ranged from 11-30%. Highest accuracy (95%CI) were found for Contec CMS50D1; 91% (86-94) and Zacurate Pro Series 500DL; 90% (85-94). The hospital-grade SpO_2_-monitor had an A_RMS_ of 3.0 and MAE of 1.9, and an accuracy of 95% (91-97%).

**Conclusion:** Direct-to-consumer pulse oximeters do not meet ISO-standards required for FDA-clearance, but can accurately rule out hypoxemia.

## BACKGROUND

Pulse oximetry has become an indispensable, low-cost, non-invasive, diagnostic tool to assess a patients’ oxygen saturation. Typically, these diagnostic instruments have a clip that can be put on a patient’s finger to obtain information on the peripheral arterial oxygen saturation (SpO_2_), which serves as a proxy for tissue oxygenation. [1] Over the past decade, pulse oximeters have evolved to play a pivotal role in routine medical care, and are an essential bedside tool in making treatment and/or referral decisions in community-based health care settings. In the past year, pulse oximeters have become even more important, as they are widely used to monitor for (silent) hypoxemia in patients with coronavirus disease 2019 (COVID-19). [2] To illustrate the uptake of pulse oximetry, the global pulse oximeter market was valued at 1.9 billion US dollars in 2019, and is predicted to grow 6.4% annually for the next decade. [3] Given its importance in guiding medical decision making, it is remarkable how little is known about the diagnostic accuracy of these devices when used under clinical conditions in actual patients. [4] As such, we aimed to evaluate whether popular direct-to-consumer fingertip pulse oximeters meet the standards for accuracy, as proposed by regulatory bodies under real-world conditions.

## METHODS

We reported this diagnostic accuracy study in accordance with the Standards for Reporting of Diagnostic Accuracy Studies (STARD) 2015 statement. [5]

### Study design

We enrolled consecutive patients, who were at least 18 years of age, admitted to the Intensive Care Unit of a large community-based hospital (Flevoziekenhuis, Almere, the Netherlands. Eligible patients had a radial artery catheter for arterial blood oxygen saturation assessments (SaO_2_) as part of routine medical care. Exclusion criteria were patients without a clinical indication for arterial access, those with inherited forms of abnormal hemoglobin, and those who rapidly deteriorated due to acute hemodynamic compromise, in which the measurements required for this study could hinder medical interventions and thereby negatively affect patient safety.

### Study procedures

Intensive Care personnel notified the site investigator (L.B.) of a potentially eligible patient for study enrolment. The site investigator enrolled each patient after verbal and written consent, either by the patient or his/her legal representative. During office hours, the site investigator was notified by the Intensive Care personnel when a blood gas sample was about to be performed. The site investigator positioned the pulse oximeters for readings directly after the arterial blood gas sample (SaO_2_) was obtained. In order to reduce detection bias the site investigator applied the pulse oximeters in a rotating fashion, maintaining a fixed device order but starting with a different device in each consecutive sample. The Intensive Care personnel (SaO_2_) and site investigator (SpO_2_ readings) reported their findings on separate digital forms, and were blinded for each other’s findings.

### Index test and devices

We evaluated ten oximeters, which were selected from the top ten of most purchased pulse oximeters on Amazon in at least two of the following countries: United States, United Kingdom, Germany, Italy, or France. Amazon was chosen because of its dominance on the e-commerce market. In alphabetical order these oximeters were: AFAC FS10D, AGPTEK FS10C, ANAPULSE ANP 100, Cocobear, Contec CMS50D1, HYLOGY MD-H37, Mommed YM101, PRCMISEMED F4PRO, PULOX PO-200, and Zacurate Pro Series 500DL. These pulse oximeters cost between 20 and 50 euros each, and all claimed to meet ISO standards (see paragraph ‘outcomes of interest’ for specifics). As a clinical index test, we also included a hospital-grade pulse oximeter (Philips M1191BL sensor glove, Philips, The Netherlands), which was used as the clinical standard of care for continuous SpO_2_ monitoring at the study site, and has met ISO standards and received 510(k) clearance of the FDA (clearance number: K062455). We used the SpO_2_ value that was shown on the pulse oximeter’s display 30 seconds after placement on a patient’s fingertip. The same fingertip was used for each oximeter. When no result was displayed 30 seconds after placement, this was documented as an invalid reading.

### Reference standard

The reference standard was a point of care testing analyser (ABL90 Flex Plus, Radiometer Medical ApS, Brønshøj, Denmark, calibrated as per regulatory standards), which was used to perform blood gas analysis on arterial blood gas samples to obtain the arterial oxygen saturation (SaO_2_) at the study site as part of routine Intensive Care.

### Outcomes of interest

We formulated the following outcome measures: mean bias, root mean square difference (A_RMS_), the mean absolute error (MAE), and diagnostic accuracy for hypoxemia, defined as SaO_2_ ≤90%. Mean bias is calculated as SpO_2_ – SaO_2_. A_RMS_ and MAE are derived from calculations involving mean bias and precision (standard deviation of bias). Because outliers have an excessive negative effect on results of the A_RMS_ parameter, we also assessed the MAE, a measurement that is more robust in the presence of outliers. The formulas to calculate these outcomes can be found in the supplement. We evaluated the diagnostic accuracy of the pulse oximeters according to the standards defined by the International Organization for Standardization in ISO 80601-2-61:2017, which supersedes the ISO 80601-2-61:2011 standard advised by the American Food and Drug Administration (FDA) in their 510(k) Premarket Notification Submissions Guidance for pulse oximeters. This standard considers an A_RMS_ of ≤3% in the SaO2 range of 70-100% acceptable. [6,7]

### Data collection and sample size

We included data on SpO_2_, SaO_2_, heart rate, systolic- and diastolic blood pressure, sex, age, skin type (FitzGerald classification) as assessed by the site investigator, noradrenalin dose (mg/h), body temperature (°C) and hand temperature to touch as assessed by the site investigator. We followed the FDA advice for sample size determination, which states that at least 200 measurements per pulse oximeter should be obtained from at least 10 subjects, of which at least 2 or 15% have a dark skin type (FitzGerald class IV-VI). [7]

### Statistical analysis

We assessed mean bias (SpO_2_ − SaO_2_) and standard deviation (SD), and subsequently calculated the A_RMS_ and MAE using the formulas as described in the supplemental data. We created Bland-Altman plots for each pulse oximeter to graphically display its bias (SpO_2_-SaO_2_) to the reference standard (SaO_2_). We added a zero-line and upper and lower limits of agreement (±1.96 SD). To visualize the accuracy standards required by the regulatory bodies, ±3% - lines are also displayed in the figures. As multiple observations were performed per individual, we calculated the standard deviation by using the within-subject variance (σ2) and the between-subject variance (σμ2). Since A_RMS_ can be easily affected by the presence of outliers, we added MAE, which is more robust in the presence of outliers, as well as a sensitivity analysis restricting the sample to measurements within 1.96 SD of each pulse oximeter’s mean and calculating A_RMS_ and MAE on this sample. This sensitivity analysis is analogous to discarding extreme readings as done in routine clinical care when a reading does not coincide with the patient’s apparent clinical state. We also assessed the diagnostic performance (sensitivity, specificity, predictive values, accuracy) of pulse oximeters in detecting hypoxemia, which we defined as SaO_2_≤90%. Finally, we evaluated factors associated with poor performance of each pulse oximeter. We used bias as a continuous measurement for performance, and used logistic regression models in which we included relevant patient and pulse oximeter characteristics. We used SPSS (IBM SPSS, version 26.0, IBM, Armonk, USA), R software (R version 3.6.1, The R Foundation for statistical computing), and MedCalc Statistical Software version 18.5 (MedCalc Software bvba, Ostend, Belgium; http://www.medcalc.org; 2018)) to conduct the analyses. We assessed statistical significance at the 0.05 level in all analyses.

### Ethics review

The study was approved by the local ethics committee and the board of directors of the Flevoziekenhuis. We obtained informed written consent from each participant. When a patient was unable to provide informed consent him-/herself, e.g. when sedated or unconscious, a legal representative provided informed consent on the patient’s behalf.

## RESULTS

### Baseline characteristics

We enrolled 35 consecutive patients, with a median age of 69 years, and 40% female. ***Table 1*** displays the baseline characteristics. Patients were primarily admitted for respiratory failure due to COVID-19 or other pulmonary diseases (such as COPD). Other diagnoses were septic shock, pulmonary embolism, and arterial thrombosis in extremities, severe hyperglycemia, and suicide attempt.

**Table 1.** Patient and pulse oximeter characteristics.

|  | Number/statistic |
| --- | --- |
| ***Patient characteristics (n=35)*** |  |
| Age, years | 69 (61-73)* |
| Female | 14 (40%) |
| Fitzpatrick IV-VI skintype | 5 (14.3%) |
| Body temperature, °C | 37.2±0.9 |
| Heart rate, beats per minute | 84 (68-101)* |
| Systolic blood pressure (mmHg) | 125 (108-143)* |
| Diastolic blood pressure (mmHg) | 57 (50-66)* |
| Noradrenaline dosage (mg/h) | 0.0 (0.0-0.3)* |
| ***Pulse oximeter characteristics (n=2,258)*** |  |
| AFAC FS10D | 232/234 (99.1%) |
| AGPTEK FS10C | 233/234 (99.6%) |
| ANAPULSE ANP 100 | 202/234 (86.3%) |
| Cocobear | 226/234 (96.6%) |
| Contec CMS50D1 | 228/234 (97.4%) |
| HYLOGY MD-H37 | 232/234 (99.1%) |
| Mommed YM101 | 229/234 (97.9%) |
| PRCMISEMED F4 PRO | 233/234 (99.6%) |
| PULOX-PO-200 | 231/234 (98.7%) |
| Zacurate Pro Series 500DL | 212/234 (90.6%) |
| Total pulse oximeter readings | 2,258/2340 (96.5%) |

Binary variables presented as number and percentage. Continuous variables were displayed as mean± standard deviation when normally distributed, or median and 25<sup>th</sup>-75<sup>th</sup> percentile (\*) when non-normally distributed.

### Blood gas samples and Oximeter readings

We obtained a total of 234 arterial blood gas samples; SaO_2_ ranged between 85.6% and 99.8%, with the distribution illustrated in ***figure 1***. Of those 234 samples, 12 samples (5.1%) were classified as hypoxemia. We obtained 2258 pulse oximeter readings, all within a 10 minute time window after acquiring the reference standard’s blood sample. Per patient we obtained on average 67 pulse oximeter measurements, ranging from 10 to 420. Of all readings, 82 (3.5%) were invalid as they did not display a result after 30 seconds. The percentage of invalid measurements varied considerably per pulse oximeter type, as shown in ***table 1***.

**Title figure 1.**
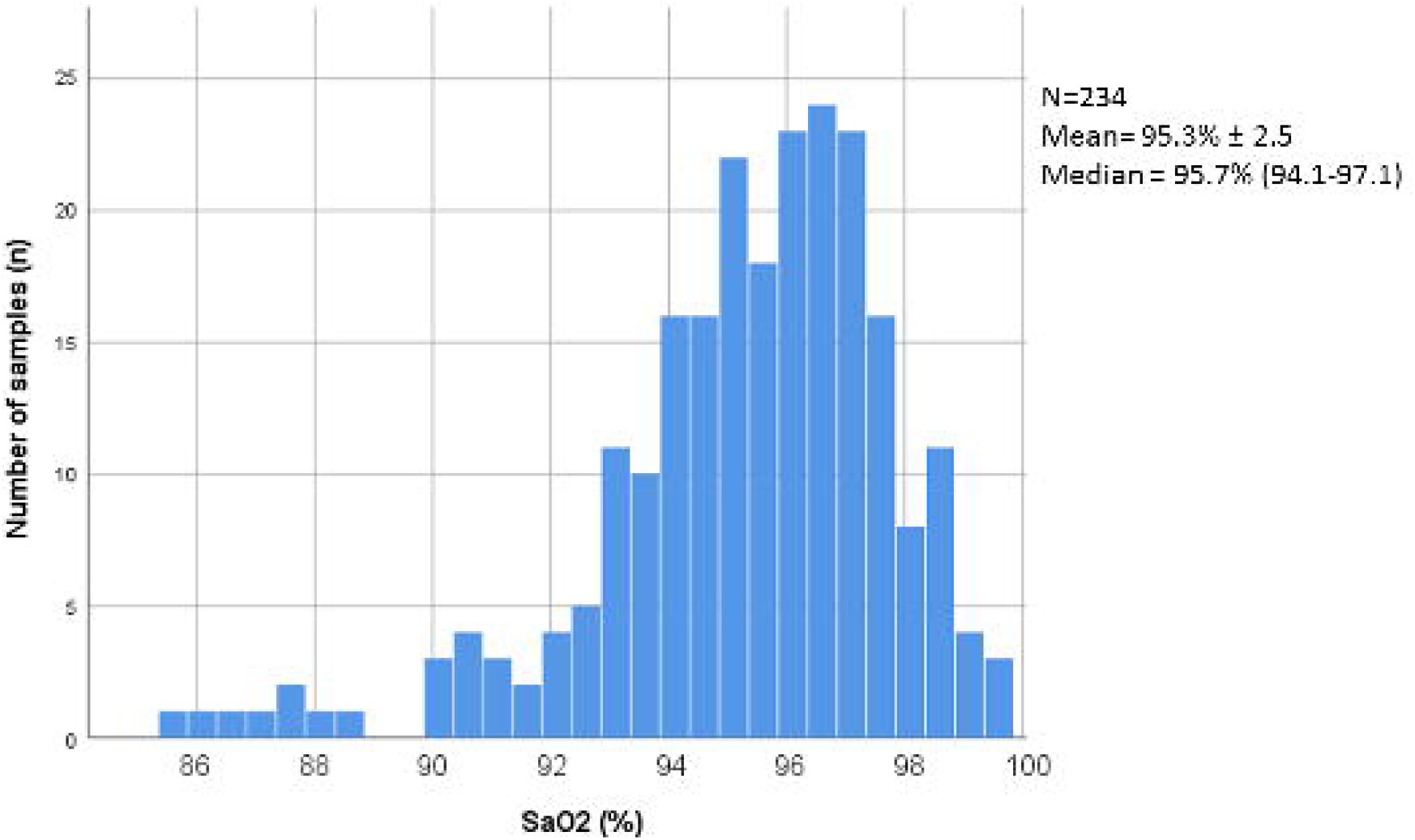
Distribution of arterial blood gas saturation (SaO_2_) among obtained samples.

### Test results of pulse oximeters

The test characteristics of the tested pulse oximeters are shown in ***table 2*** and are graphically displayed in ***figure 2*** and ***figure S1***. We found that most pulse oximeters overall had a negative mean bias, thus SpO_2_ was on average lower than SaO_2_. Moreover, none of the tested pulse oximeters met the requirement of diagnostic accuracy, using the A_RMS_ ≤3% threshold. When using MAE, which is less affected by outliers, five oximeters performed within a ≤3% difference margin of the reference standard. When excluding extreme outliers (>1.96 SD), performance of Contec CMS50D1, Mommed YM101, Pulox-PO-200, and Zacurate Pro Series 500DL were within the A_RMS_ ≤3% limit, and all but two oximeters (ANAPULSE ANP 100 and Cocobear) met the MAE ≤3% thresholds.

**Table 2.** Performance of pulse oximeters compared with arterial blood gas (using the metrics: mean bias, root mean square difference and mean absolute error)

|  | All measurements including outliers |  |  | Excluding outliers* |  |
| --- | --- | --- | --- | --- | --- |
| | Mean bias (±SD) | A<sub>RMS</sub><sup>α</sup> (%) | MAE<sup>β</sup> (%) | A<sub>RMS</sub><sup>α</sup> (%) | MAE<sup>β</sup> (%) |
| AFAC FS10D | -2.6 ± 4.4 | 5.1 | 3.1 | 3.3 | 2.5 |
| AGPTEK FS10C | -2.2 ± 4.0 | 4.6 | 2.9 | 3.0 | 2.3 |
| ANAPULSE ANP 100 | -4.8 ± 5.9 | 7.5 | 5.1 | 5.4 | 4.4 |
| Cocobear | -3.7 ± 5.1 | 6.2 | 4.0 | 4.3 | 3.4 |
| Contec CMS50D1 | -0.6 ± 4.5 | 4.5 | 2.3 | 2.7 | 2.0 |
| HYLOGY MD-H37 | -3.1 ± 4.9 | 5.7 | 3.5 | 3.6 | 2.8 |
| Mommed YM101 | -1.7 ± 3.6 | 3.9 | 2.4 | 2.6 | 1.9 |
| PRCMISEMED F4 PRO | -2.4 ± 6.4 | 6.8 | 3.5 | 3.5 | 2.6 |
| PULOX-PO-200 | -1.6 ± 3.8 | 4.1 | 2.6 | 2.8 | 2.2 |
| Zacurate Pro Series 500DL | -1.4 ± 4.5 | 4.7 | 2.5 | 2.5 | 1.9 |

\* only values within the limits of agreement (within 1.96 standard deviation)
<sup>$\alpha$</sup> A<sub>RMS</sub>: root mean square difference
<sup>$\beta$</sup> MAE: mean absolute error

**Title figure 2.**
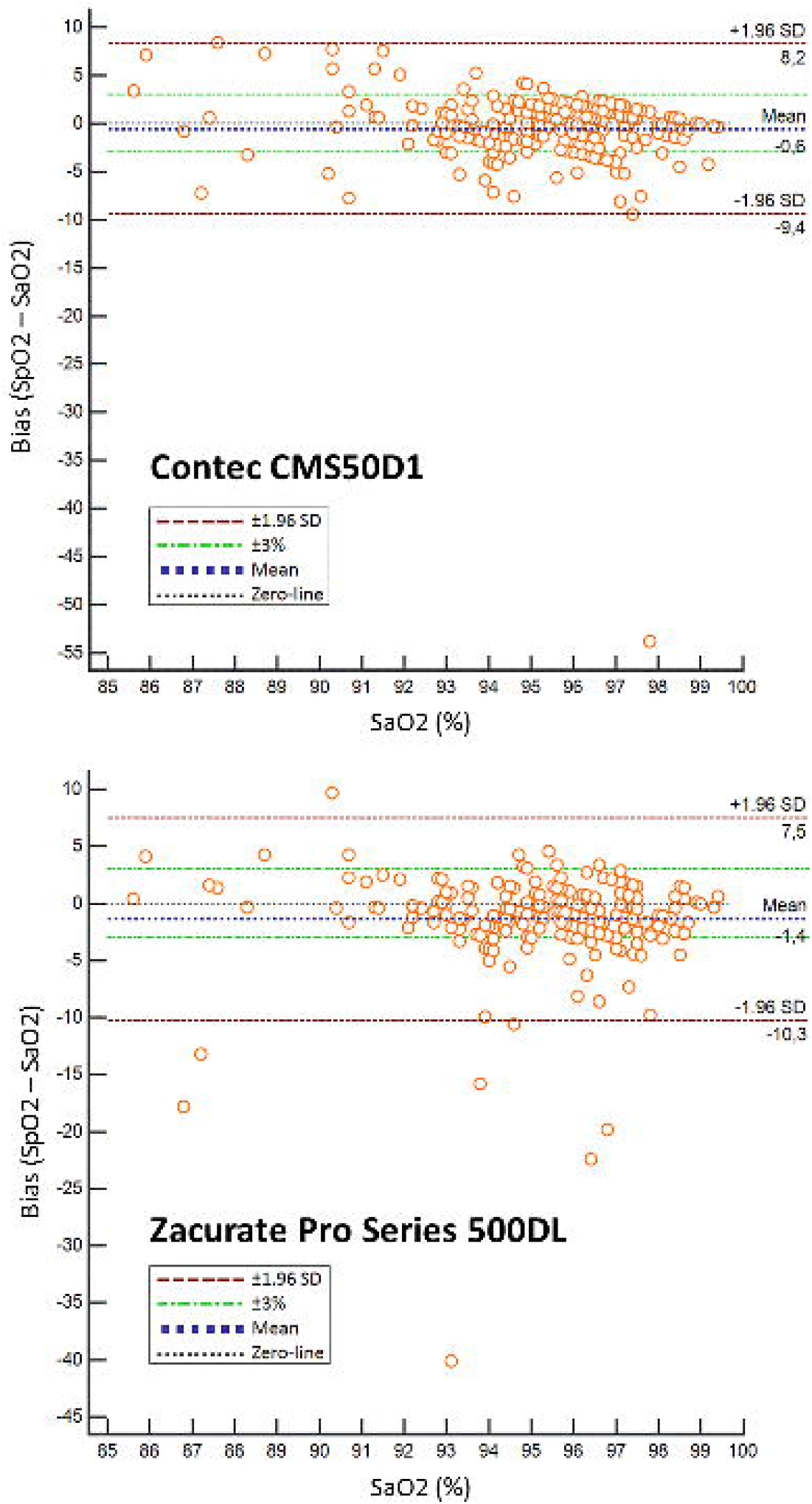
Bland-Altman plots of the bias compared with SaO_2_ of two pulse oximeters with the lowest mean bias (Contec CMS50D1 −0.6 and Zacurate Pro series 500DL −1.4).* **Legend figure 2:** Bland-Altman plots for each pulse oximeter to graphically display its bias (SpO_2_-SaO_2_) to the reference standard (SaO_2_). We added a zero-line and upper and lower limits of agreement (±1.96 SD). To visualize the accuracy standards required by the regulatory bodies, ±3% - lines are also displayed in the figures. *Bland-Altman plots of the other pulse oximeters can be found as a supplemental figure.

### Detection of hypoxemia

***Table 3*** summarizes the accuracy of each pulse oximeter in detecting hypoxemia. Overall, the oximeters with the highest specificity were Contec CMS50D1 (93%) and Zacurate Pro Series 500DL (91%). Oximeters with the highest sensitivity were Hylogy MD-H37 (92%) and Anapulse ANP 100 (91%). In terms of predictive values, all pulse oximeters performed well in ruling out hypoxemia, with negative predictive values of 98-99% in a population with ≈5% hypoxemia measurements. Confirming the presence of hypoxemia was poor, with positive predictive values of 11 to 30%. For all reading, accuracy was highest for Contec CMS50D1 (91%) and Zacurate Pro Series 500DL (90%). As a sensitivity analysis, we also provided data on the performance of pulse oximeters at different abnormal oxygenation thresholds (SaO_2_≤92% and SaO_2_≤94), which can be found as supplemental ***table S1***.

**Table 3.** Accuracy of pulse oximeters in detecting hypoxemia (prevalence of SaO_2_≤90% is 5.1%)

| Pulse oximeter | TP/FN | FP/TN | Sensitivity | Specificity | PPV | NPV | Accuracy |
| --- | --- | --- | --- | --- | --- | --- | --- |
| AFAC FS10D | 9/3 | 46/174 | 75 (43-95) | 79 (73-84) | 16 (11-23) | 98 (96-99) | 79 (73-84) |
| AGPTEK FS10C | 10/2 | 39/182 | 83 (52-98) | 82 (77-87) | 20 (15-27) | 99 (96-100) | 82 (77-87) |
| ANAPULSE ANP 100 | 10/1 | 79/112 | 91 (59-100) | 59 (51-66) | 11 (9-14) | 99 (95-100) | 60 (53-67) |
| Cocobear | 10/2 | 63/151 | 83 (52-98) | 71 (64-77) | 14 (10-18) | 99 (96-100) | 71 (65-77) |
| Contec CMS50D1 | 7/5 | 16/200 | 58 (28-85) | 93 (88-96) | 30 (18-46) | 98 (95-99) | 91 (86-94) |
| HYLOGY MD-H37 | 11/1 | 51/169 | 92 (62-100) | 77 (71-82) | 18 (14-22) | 99 (96-100) | 78 (72-83) |
| Mommed YM101 | 9/3 | 31/186 | 75 (43-95) | 86 (80-90) | 23 (15-32) | 98 (96-99) | 85 (80-89) |
| PRCMISEMED F4 PRO | 8/4 | 39/182 | 67 (35-90) | 82 (77-87) | 17 (11-25) | 98 (95-99) | 82 (76-86) |
| PULOX-PO-200 | 9/3 | 30/189 | 75 (43-95) | 86 (81-91) | 23 (16-32) | 98 (96-99) | 86 (81-90) |
| Zacurate Pro Series 500DL | 8/2 | 19/183 | 80 (44-97) | 91 (86-94) | 30 (20-42) | 99 (96-100) | 90 (85-94) |

The values for PPV, NPV and accuracy are dependent on disease prevalence
TP= true positive, FP= false positive, FN= false negative, TN= true negative, PPV= positive predictive value, NPV= negative predictive value.

### Factors associated with poor pulse oximeter performance

***Table 4*** displays factors that were associated with bias (SpO_2_-SaO_2_) of each pulse oximeter. Overall, darker skin complexion and an inaccurate pulse rate measurement (difference between pulse oximeter pulse rate and “true” pulse rate as captured by ICU monitor), were associated with poorer SpO_2_ performance in five out of ten pulse oximeters. Other factors that negatively affected results were increased systolic blood pressure and cold hands to touch, in four and three pulse oximeters, respectively.

**Table 4.**
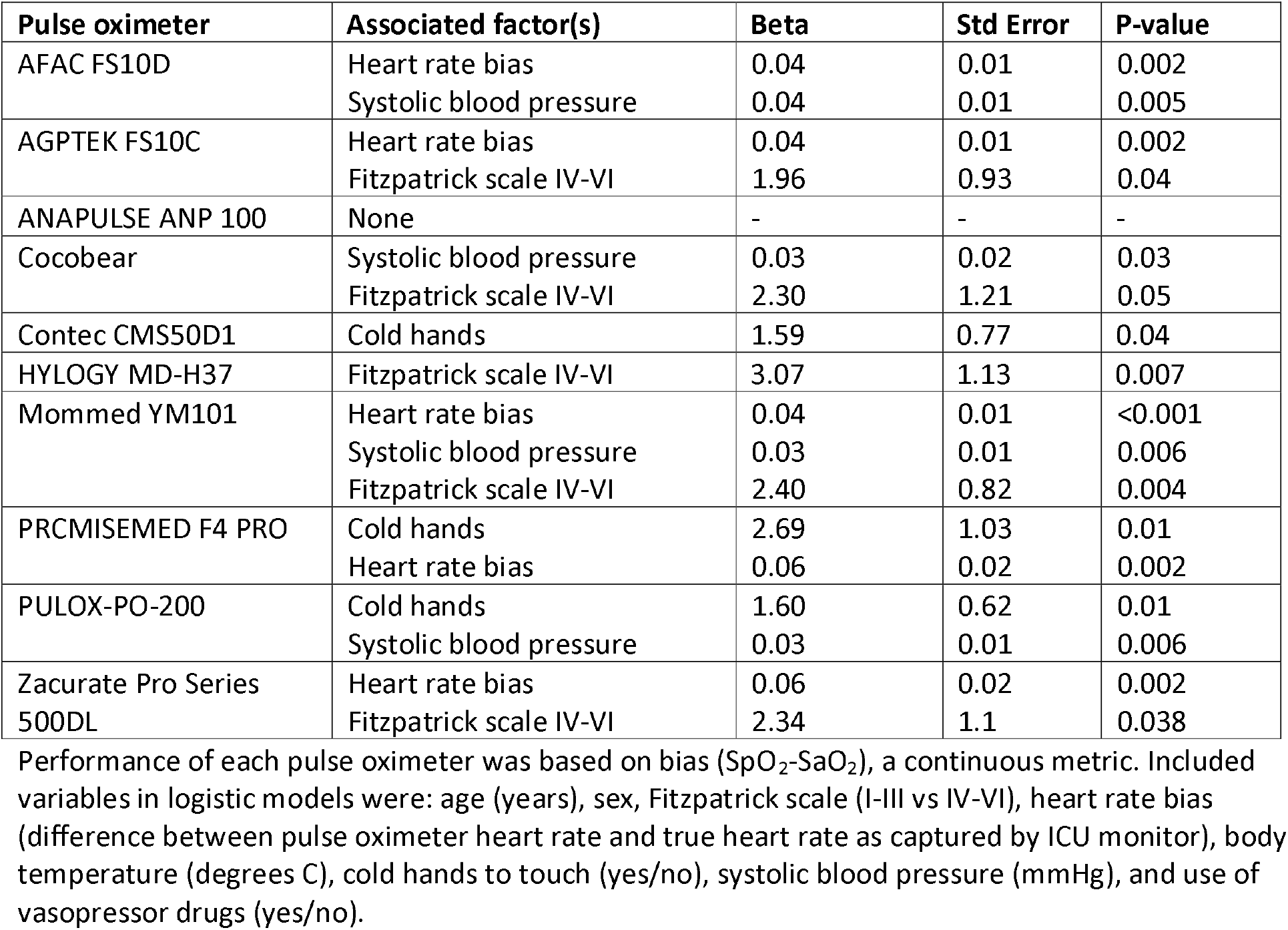
Factors associated with poor performance of each pulse oximeter.

### Test results and diagnostic accuracy of the SpO_2_ continuous monitor (clinical index test)

The hospital-grade Philips sensor glove SpO_2_ continuous monitor performed with an A_RMS_ of 3.0 and MAE of 1.9 for all measurements, and A_RMS_ of 2.1 and MAE of 1.6 when excluding outliers (1.96 SD). For detecting hypoxemia, the hospital grade Philips sensor glove SpO_2_ monitor had a sensitivity of 67% (35-90%), specificity of 96% (93-98%), positive and negative predictive values of 50% (31-69) and 98% (96-99%), respectively, and an accuracy of 95% (91-97%). The presence of cold hands/acra was associated with higher inaccuracy of the SpO_2_ continuous monitor compared with SaO_2_.

## DISCUSSION

Our study found that the top ten of low-cost, popular direct-to-consumer pulse oximeters do not meet the requirements set by the regulatory bodies (ISO/FDA), when compared with the gold standard of arterial oxygen saturation, as obtained by blood gas samples, in a population of intensive care patients. However, when extreme outliers were disregarded, four pulse oximeters would meet the A_RMS_≤3% requirements, and eight pulse oximeters would meet MAE≤3% standards. The hospital grade Philips sensor glove SpO_2_ monitor performed slightly better. From a clinical perspective, we found that the pulse oximeters perform well in ruling out hypoxemia, but are not reliable in detecting manifest hypoxemia. As such, when a pulse oximeter indicates a low SpO_2_, confirmation is required. Caution is warranted when factors are present that negatively affect the reliability of pulse oximeters, such as an inaccurate pulse rate reading, darker skin pigmentation, high blood pressure, or cold extremities.

### Strengths and limitations

Our study was performed under clinical conditions involving consecutive patients with direct access to arterial blood gas samples. Patients were diverse in terms of age, skin type, and underlying clinical conditions. We used popular pulse oximeter devices and the number of samples was sufficient to draw accurate conclusions. Moreover, we used a hospital grade SpO_2_ device as a clinical index test. However, there are also a number of limitations that should be mentioned. First, the performance of fingertip pulse oximeters may have been affected by the poor health status and poorer peripheral perfusion of intensive care patients, when compared with community-based patients. Moreover, the distribution of SaO_2_ is different in intensive care populations (i.e. oxygenation and ventilation settings are set to strive for SaO_2_ of 92%), when compared with community based populations, which would also affect performance. Finally, we did not capture detailed laboratory findings on our patients, such as hemoglobin or acid-base status. These variables are of relevance as SpO_2_ readings are based on photopletysmographic measurements using infrared wavelengths through (de)oxyhemoglobin. As such, a decrease in hemoglobin blood concentration may affect capturing a sufficient signal, whereas a pH shift could alter the oxyhemoglobin dissociation curve, both resulting in inaccurate SpO_2_ calculations.

### Prior studies

Many commonly used direct-to-consumer pulse oximeters do not undergo rigorous in vivo testing, and thus, little is known about the accuracy of these devices. An important study which did perform in vivo testing of inexpensive pulse oximeters was published by Lipnick *et al* in 2016. [4] In this study six finger pulse oximeters (not cleared by the FDA) were evaluated in 22 healthy subjects, in which stable SaO_2_ plateaus between 70-100% were achieved under controlled conditions via a partial rebreathing circuit. The study found that two pulse oximeters tested (Contec CMS50DL and Beijing Choice C20) demonstrated an A_RMS_ of ≤3%, hereby meeting the ISO criteria for accuracy. Of the tested oximeters, the Contec CMS50DL may be comparable with the Contec CMS50D1 model that we tested. In our study, which was performed under clinical conditions, the Contec device did not fare as well, although it was still one of the better pulse oximeters that we tested. Prior studies found that oximeters performed worse in hypoxic conditions, with mean bias increasing at lower oxygen saturations compared with arterial blood gas [4] or a conventional bedside pulse oximeter [8,9] In our study we found a similar observation in some, but not all oximeters. Still, from a clinical perspective, it is particularly important to have minimal bias in the range of 90 to 95%, as this is where a hypoxemic state should be differentiated from a non-hypoxemic state.

### Implications for practice

Due to the current COVID-19 pandemic, pulse oximeters have become more popular than ever before. Despite their limitations, these devices present a welcome tool for remote monitoring of patients and for ruling out hypoxemia, particularly in a population where the prevalence of hypoxemia is low. In our population, in which approximately 5% was hypoxemic, these low-cost devices were able to safely rule out hypoxemia in virtually all cases (98-99%). This percentage would even further approach 100% in low prevalence settings. Still, our findings, as those of other studies [4,8,9] illustrate that in patients with other symptoms suggestive of hypoxemia, physicians should remain alert, particularly in high-risk patients with preexisting pulmonary disease. In these scenarios, devices that are FDA-cleared, should be preferred as they show a smaller degree of increasing bias during lower SaO_2_ conditions. [10] Finally, irrespective of the device used, it is important for clinicians to realize that there are a number of factors that negatively impact the reliability of pulse oximetry. The most important ones include anemia, nail polish, and dark skin pigmentation [11,12], as well as others such as poor peripheral circulation, and inaccurate pulse rate measurement, as shown in our study.

## CONCLUSION

Low-cost pulse oximeters are widely available and in high demand. Most pulse oximeters have not been rigorously tested. In this study, we tested ten popular pulse oximeters in ICU patients with direct access to arterial blood gas. Overall, we found that the tested pulse oximeters would not meet strict ISO requirements used by the FDA in their 510(k) premarket notification clearance process. However, most devices can safely rule out hypoxemia in the vast majority of patients, particularly in community-based populations with a lower *a priori* hypoxemia risk. Future studies are warranted to further assess the accuracy of pulse oximeters in community-based patients, and to gain insight how to further improve this non-invasive, low-cost, and potentially life-saving technology.

## Data Availability

All data relevant to the study are included in the article or uploaded as an online supplement. The datasets used during the current study are available from the corresponding author on reasonable request.

## Contributors

RH and LB designed the study protocol. LB was involved in the recruitment of patients for participation in data collection, supervised by MS. LB performed the statistical analysis and interpreted all data with statistical guidance from JH, LdC, EK, WL, and RH. RH and LB drafted the manuscript and all authors contributed to its revision. All authors read and approved the final manuscript

## Acknowledgements

We would like to thank all participating patients, their families, as well as the personnel of the Intensive Care of FlevoZiekenhuis.

## Funding

This research received no specific grant from any funding agency in the public, commercial or not-for-profit sectors.

## Competing interests

The authors report no competing interests with regards to the current study.

## Supplemental data

### Definition of Root mean square difference

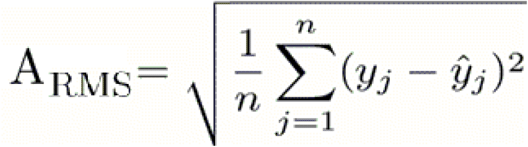

*A*_*RMS*_: *root mean square difference;*

*n: number of samples;*

*y*_*j*_: *Pulse oximeter measurement;*

*ŷ* _*j*_: *Reference standard measurement*

We squared the difference (either positive, negative or 0) between a pulse oximeter measurement and the reference standard measurement. Then we calculated the mean of the sum of squares, and determined the root of that mean. As the difference between the pulse oximeter and the reference standard is squared, the A_RMS_ is always positive.

### Definition of Mean absolute error

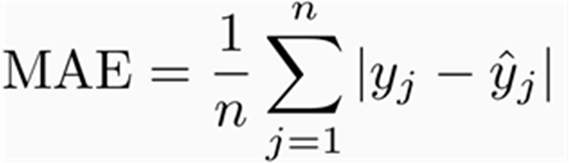

*MAE: mean absolute error;*

*n: number of samples;*

*y* _*j*_: *Pulse oximeter measurement;*

*ŷ* _*j*_: *Reference standard measurement*

We calculated the absolute difference (either positive or 0) between a pulse oximeter measurement and the reference standard measurement. We then determined the mean of these absolute differences. Similarly to the A_RMS_, the MAE is always positive, as it concerns an absolute difference.

---

**Table S1.** Accuracy of pulse oximeters in detecting abnormal oxygenation status, using either SaO_2_≤92% or SaO_2_≤94% as threshold.

| **$\text{SaO}_2 \leq 92\%$** | **Pulse oximeter** | **TP/FN** | **FP/TN** | **Sensitivity** | **Specificity** | **PPV** | **NPV** | **Accuracy** |
| --- | --- | --- | --- | --- | --- | --- | --- | --- |
|  | AFAC FS10D | 19/6 | 66/141 | 76 (55-91) | 68 (61-74) | 22 (18-28) | 96 (92-98) | 69 (63-75) |
|  | AGPTEK FS10C | 18/7 | 64/144 | 72 (51-88) | 69 (63-75) | 22 (17-28) | 95 (92-97) | 70 (63-75) |
|  | ANAPULSE ANP 100 | 18/5 | 76/103 | 78 (56-93) | 58 (50-65) | 19 (15-24) | 95 (90-98) | 60 (53-67) |
|  | Cocobear | 22/1 | 90/113 | 96 (78-100) | 56 (49-63) | 20 (17-23) | 99 (94-100) | 60 (53-66) |
|  | Contec CMS50D1 | 13/12 | 34/169 | 52 (31-72) | 83 (77-88) | 28 (19-38) | 93 (90-96) | 80 (74-85) |
|  | HYLOGY MD-H37 | 21/4 | 79/128 | 84 (64-95) | 62 (55-68) | 21 (17-25) | 97 (93-99) | 64 (58-70) |
|  | Mommed YM101 | 18/7 | 52/152 | 72 (51-88) | 75 (68-80) | 26 (20-33) | 96 (92-98) | 74 (68-80) |
|  | PRCMISEMED F4 PRO | 15/9 | 64/145 | 63 (41-81) | 69 (63-76) | 19 (14-25) | 94 (91-96) | 69 (62-75) |
|  | PULOX-PO-200 | 20/5 | 52/154 | 80 (59-93) | 75 (68-81) | 28 (22-34) | 97 (93-99) | 75 (69-81) |
|  | Zacurate Pro Series 500DL | 15/7 | 37/153 | 68 (45-86) | 81 (74-86) | 29 (21-38) | 96 (92-98) | 79 (73-85) |
| **$\text{SaO}_2 \leq 94\%$** |  |  |  |  |  |  |  |  |
|  | AFAC FS10D | 64/6 | 76/86 | 91 (82-97) | 53 (45-61) | 46 (41-50) | 93 (87-97) | 65 (58-71) |
|  | AGPTEK FS10C | 62/8 | 71/92 | 89 (79-95) | 56 (48-64) | 47 (42-51) | 92 (86-96) | 66 (60-72) |
|  | ANAPULSE ANP 100 | 60/5 | 102/35 | 92 (83-97) | 26 (18-34) | 37 (34-40) | 88 (74-94) | 47 (40-54) |
|  | Cocobear | 71/2 | 88/71 | 97 (90-100) | 45 (37-53) | 45 (41-48) | 97 (90-99) | 61 (55-68) |
|  | Contec CMS50D1 | 55/15 | 41/117 | 79 (67-87) | 74 (66-81) | 57 (50-64) | 89 (83-93) | 75 (69-81) |
|  | HYLOGY MD-H37 | 67/3 | 86/76 | 96 (88-99) | 47 (39-55) | 44 (40-48) | 96 (89-99) | 62 (55-68) |
|  | Mommed YM101 | 60/9 | 56/104 | 87 (77-94) | 65 (57-72) | 52 (46-57) | 92 (86-96) | 72 (65-77) |
|  | PRCMISEMED F4 PRO | 57/12 | 75/89 | 83 (72-91) | 54 (46-62) | 43 (38-48) | 88 (81-93) | 63 (56-69) |
|  | PULOX-PO-200 | 62/7 | 70/92 | 90 (80-96) | 57 (49-65) | 47 (42-52) | 93 (87-96) | 67 (60-73) |
|  | Zacurate Pro Series 500DL | 55/9 | 45/103 | 86 (75-93) | 70 (62-77) | 55 (48-61) | 92 (86-95) | 75 (68-80) |

The values for PPV, NPV and accuracy are dependent on disease prevalence
TP= true positive, FP= false positive, FN= false negative, TN= true negative, PPV= positive predictive value, NPV= negative predictive value.

**Figure S1.**
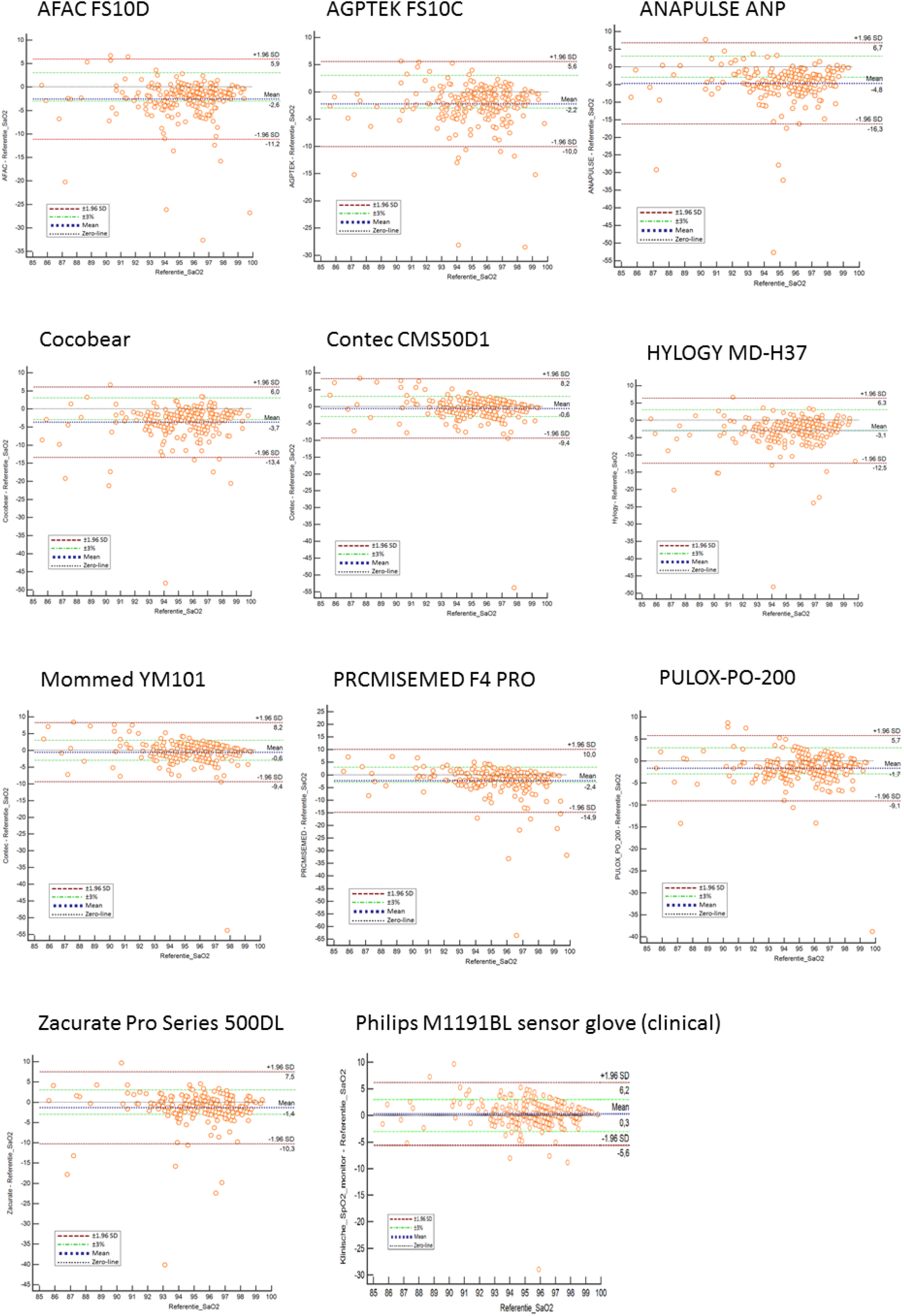
Bland-Altman plots of performance of pulse oximeters (bias compared with SaO_2_.

## Notes

### Competing Interest Statement

The authors have declared no competing interest.

### Clinical Trial

This study did not involve a clinical trial

### Author Declarations

The study was approved by the local ethics committee and the board of directors of the Flevoziekenhuis

## REFERENCES

[1] Jubran A. Pulse oximetry. Crit Car 2015;19:272.

[2] Tobin, M., F. Laghi, and A. Jubran, Why COVID-19 Silent Hypoxemia Is Baffling to Physicians. American Journal of Respiratory and Critical Care Medicine, 2020. 202(3): p. 356–360.

[3] Grand View Research. Pulse Oximeters Market Size, Share & Trends Analysis Report By Type (Fingertip, Handheld), By End Use (Hospitals & Healthcare Facilities, Homecare), By Region (North America, Europe, APAC, LATAM, MEA), And Segment Forecasts, 2020 – 2027. Pub June 2020. https://www.grandviewresearch.com/industry-analysis/pulse-oximeter-market/toc

[4] Lipnick MS, Feiner JR, Au P, Bernstein M, Bickler PE. The accuracy of 6 inexpensive pulse oximeters not cleared by the Food and Drug Administration: the possible global public health implications. Anesth Analg 2016;123:338–45.

[5] Bossuyt PM, Reitsma JB, Bruns DE, Gatsonis CA, Glasziou PP, Irwig L, LijmerJG Moher D, Rennie D, de Vet Hcw, Kressel HY, Rifai N, Golub RM, Altman DG, Hooft L, Korevaar DA, Cohen JF, For the STARD Group. STARD 2015: An Updated List of Essential Items for Reporting Diagnostic Accuracy Studies. BMJ 2015;351:h5527.

[6] Medical electrical equipment — Part 2-61: Particular requirements for basic safety and essential performance of pulse oximeter equipment. 2017, International Organization for Standardization.

[7] Pulse Oximeters - Premarket Notification Submissions [510(k)s] Guidance for Industry and Food and Drug Administration Staff, U.S.D.o.H.a.H. Services, Editor. 2013, U.S. Department of Health and Human Services: Rockville. p. 7.

[8] Smith RN, Hofmeyr R. Perioperative comparison of the agreement between a portable fingertip pulse oximeters v. a conventional bedside pulse oximeter in adult patients (COMFORT trial). South Afr Med J 2019;3:109:154–158

[9] Hudson AJ, Benjamin J, Jardeleza T, Bergstrom C, Cronin W, Mendoza M, Schultheis L. Clinical interpretation of peripheral pulse oximeters labeled “not for medical use”. Ann Fam Med 2018;16:552–554.

[10] Luks AM, Swenson ER. Pulse oximetry for monitoring patients with COVID-19 at home. Potential pitfalls and practical guidance. Ann Am Thorac Soc 2020;17:1040–1046.

[11] Adler JN, Hughes LA, Vivilecchia R, Camargo CA Jr. Effect of skin pigmentation on pulse oximetry accuracy in the emergencydepartmen. Acad Emerg Med 1998;5:965–70.

[12] Coté CJ, Goldstein EA, Fuchsman WH, Hoaglin DC. The effect of nail polish on pulse oximetry. Anesth Analg 1988;67:683–6

## References

[1] Kampakis, S. Performance measures RMSE MAE. 2020 [cited 2020 01-10-2020]; Available from: https://thedatascientist.com/performance-measures-rmse-mae/.

[2] Lipnick;, M.S., The Accuracy of 6 Inexpensive Pulse Oximeters Not Cleared by the Food and Drug Administration: The Possible Global Public Health Implications. Anesthesia and Analgesia, 2016. 123(2): p. 339–344.

